# Effects of Immediate Postpartum Diuretic Treatment on Postpartum Blood Pressure among Individuals with Hypertensive Disorders of Pregnancy: A Systematic Review and Meta-Analysis

**DOI:** 10.1101/2025.01.03.25319983

**Authors:** Susan K. Keen, Koura Sall, Agnes Koczo, Yisi Wang, Rebekah S. Miller, Matthew F. Muldoon, Alisse K. Hauspurg, Malamo E. Countouris

## Abstract

**Background:** Hypertensive disorders of pregnancy (HDP) are associated with ongoing postpartum hypertension (HTN) and increased morbidity. Extravascular water and sodium mobilization is implicated in postpartum blood pressure (BP) elevation, however trials of postpartum diuretics in HDP have had mixed results. Our meta-analysis aimed to analyze the impact of postpartum diuretics on postpartum hypertension following HDP.

**Methods:** Systematic review identified randomized controlled trials (RCTs) studying the efficacy of diuretics in the treatment of postpartum BP. Meta-analysis outcomes included persistent HTN up to 10 days postpartum, mean postpartum systolic and diastolic BPs, and use of additional antihypertensive medications.

**Results:** From 9 RCTs, 1273 subjects were included in the meta-analysis. Postpartum diuretic use was associated with lower systolic BP (SMD standard mean difference]: -0.36; 95% confidence interval [CI]: -0.72; -0.01) without a difference in diastolic BP (SMD: 0.01; 95% CI: -0.22; 0.23) compared with controls. There was no difference in rates of persistent HTN between the postpartum diuretics group versus controls (OR: 0.70; 95% CI: 0.4; 1.05) or in antihypertensive medication use (OR: 0.66; 95% CI: 0.42; 1.05).

**Conclusion:** Postpartum diuretic use was associated with lower systolic BP compared with controls and non-significant trends of lower rates of persistent HTN and postpartum antihypertensive medication use. Due to the low certainty of evidence, uniform postpartum diuretic use with HDP cannot be recommended. Future studies are needed to evaluate specific HDP subgroups who may benefit from diuretic use.

## INTRODUCTION

Hypertensive disorders of pregnancy (HDP) impact 10-20% of pregnant individuals in the United States and are associated with increased morbidity in the peripartum period including increased risk of hospital readmission, severe hypertension (HTN), heart failure, and stroke.^1–4^ Preeclampsia and gestational HTN comprise the majority of these cases, while chronic HTN and superimposed preeclampsia account for the remainder.^5^ Postpartum HTN most commonly occurs among individuals with antenatally diagnosed HDP but can also be due to the development of a de novo postpartum process.^4^

Normal pregnancy is characterized by a 40-50% expansion of the circulating blood volume which is associated with retention of sodium and water in the interstitial tissues and a gradual rise in systolic and diastolic blood pressure (BP) towards the end of pregnancy.^6, 7^ BP rises after delivery, peaking at 3 to 10 days postpartum.^8, 9^ This postpartum BP elevation has been attributed to extravascular water and sodium mobilization into the intravascular space and may be compounded by iatrogenic administration of intravenous fluid during labor or cesarean section.^10^ Further, signs and symptoms of volume overload are prevalent among individuals with HDP, including dyspnea (20-30%), pulmonary edema (11%), and peripheral edema (11-18%).^4^ Therefore, diuretics have been proposed to theoretically accelerate postpartum BP recovery among individuals with HDP through urinary sodium and water excretion, thereby decreasing intravascular volume.^11, 12^ Interventions that improve postpartum BP may reduce associated maternal morbidity.

While several clinical guidelines have put forth recommendations for BP monitoring in the postpartum period, namely, checking an ambulatory BP within 10 days of delivery, there is a paucity of specific recommendations for postpartum BP management. According to the American College of Obstetricians and Gynecologists, treatment of postpartum HTN is recommended for systolic BP ≥150 mm Hg or diastolic BP ≥100 mm Hg with the choice of medication primarily limited by compatibility with breastfeeding and use of contraception.^5^ Current guidance on the management of HDP focuses on the use of calcium channel blockers and nonselective beta blockers but does not include specific recommendations for diuretic use. As a result, diuretic use is typically reserved for postpartum management of pulmonary edema or systemic volume overload. Given a proposed pathophysiology of HDP through intravascular volume expansion and increased sodium body sodium content, there is an opportunity to improve BP control with diuretics.

Randomized controlled trials evaluating the effectiveness of diuretic treatment for individuals with HDP in the postpartum period have been small. Further, studies have reported mixed results regarding BP effects. The latest meta-analysis found postpartum loop diuretics was associated with no difference in persistent hypertension or need for antihypertensive therapy at discharge.^7^ With the recent publication of new, relevant studies as well as the lack of inclusion of all diuretic classes in prior meta-analyses, there is a need for a contemporary investigation on the role of diuretics in the postpartum setting after HDP. In this study, we performed an updated systematic review and meta-analysis to compare the effect of immediate postpartum diuretic treatment to placebo or alternative therapies on reducing persistent postpartum HTN, systolic and diastolic BP, need for additional antihypertensive medication, and postpartum complications among individuals with HDP.

## METHODS

### Data Sources and Searches

We registered the protocol for this review in the PROSPERO database (no. CRD42021253740). Searches were developed by a health sciences librarian and performed in the Ovid Medline, Embase.com, Web of Science Core Collection, and Cochrane CENTRAL databases (see Supplemental Appendix for full search strings). The search strings included natural language and database-specific controlled vocabulary covering the concepts HDP and diuretic medications. The searches were limited to the English language as well as the years 1980 to present. Additionally, clinicaltrials.gov was searched for relevant trials. On May 11, 2021, 3718 references were downloaded into the EndNote reference management software. The references were deduplicated twice: first using the Amsterdam Efficient Deduplication method and then the Bramer et al. method, resulting in a total of 2994 citations.^13, 14^ These citations were uploaded to the DistillerSR software (Distiller, Evidence Partners, Ottawa, Canada) for screening. To supplement these electronic searches, reference lists of pertinent articles were reviewed to identify any additional potential eligible studies. Ongoing surveillance was conducted using PubMed article alerts to identify any potential additional studies published through December 1, 2024. The search was rerun on June 29, 2022, and after deduplication of the search, 246 new citations were uploaded to Distiller.^15^

### Study Selection

In Distiller, two of four cardiology or obstetric physician investigators (SK, MC, AK, and AH) independently reviewed each title, abstract, and full text article when needed using prespecified inclusion and exclusion criteria (Supplemental Table 1). Disagreements were resolved by discussion. Randomized controlled trials (RCTs) and observational cohort studies (prospective or retrospective) were eligible for inclusion. Case control studies, cross-sectional studies, case reports, conference presentations or posters, and narrative review articles were excluded. Eligible interventions included intravenous or oral diuretics or diuretics in conjunction with another antihypertensive pharmacotherapy, while eligible comparators included placebo or non-diuretic antihypertensive pharmacotherapy. Primary outcomes included “persistent HTN” defined as two BP readings taken four hours apart with systolic BP ≥ 140 mmHg or diastolic BP ≥ 90 mmHg before delivery hospitalization discharge (day 0) or up to 10 days postpartum. As guidelines recommend checking an ambulatory BP within 10 days of delivery in individuals with HDP, we performed our analysis by grouping persistent HTN from 0-10 days postpartum.

Additional outcomes include systolic and diastolic BP, persistent HTN at 30 days or 6 weeks postpartum, need for additional antihypertensive medication, length of hospitalization, hospital readmission, emergency department visits, heart failure, b-type natriuretic peptide levels, and lactation outcomes.

### Data Extraction and Quality Assessment

For each included study, one of four reviewers (SK, MC, AK, or KS) abstracted relevant study characteristics into a structured form. A second reviewer (SK, MC, AK, or KS) verified all data for accuracy. For each included study, two reviewers (SK, MC, AK, or KS) independently assessed methodological quality. As only RCT studies reached the full text screening stage, the Cochrane collaboration’s risk-of-bias tool for randomized trials version 2-0 was used.^16^ This tool allowed reviewers to assess bias in 5 categories including bias arising from the randomization process, bias due to deviations from intended interventions, bias due to missing outcome data, bias in measurement of the outcome, and bias in selection of the reported result. Through a series of yes/no questions, a risk of bias score (low, some concerns, high) was assigned per domain for each study. A low risk of bias meant that the study was judged to be a low risk of bias in all domains. A “some concern” risk of bias meant that the study was judged to have risk of concerns in some domains but no high risk of bias in any domains. Finally, a high risk of bias meant that the study was judged to have at least one high risk domain or multiple domains with some concern of bias overall lowering the confidence of the results. In turn, these domain-level judgements provided the basis for an overall risk-of-bias judgment for the specific trial result being assessed through an algorithm built in the tool. At the end of this thorough review, two reviewers (SK and KS) compared risk of bias scores per trial and any disagreements in data extraction or study quality assessments were resolved by consensus or with a third reviewer.

### Statistical Analyses

Bias corrected standardized mean differences (Hedges’ g) were used to present the difference between outcomes. A random effects model was used to estimate a pooled relative risk for persistent postpartum HTN. A Forest plot was used to illustrate the individual and pooled relative risk, estimate the confidence interval for the pooled relative risk, and report an overall p- value. Secondary continuous outcomes were assessed using a random effects model to compute a pooled mean difference. All statistical tests were performed in R 4.4.1 (R version 4.4.1 (2024-06-14 ucrt).^17^ A pooled treatment effect model (theta) p-value <0.05 was considered significant.

## RESULTS

### Characteristics of Included Studies

The searches yielded 3973 citations as summarized in the PRISMA flow diagram (Figure 1). After removing duplicates, 3240 unique studies were screened and 95 were sought for retrieval. Out of these 95 studies, 86 were excluded due to various ineligibilities including study design, population, timing of intervention, comparison, outcomes, interventions, and additional duplicate records. In addition, one recently concluded clinical trial (DIUPRE) registered on clinicaltrials.gov met inclusion criteria. Unpublished study results were obtained from trial investigators and included in the meta-analysis.

**Figure 1.**
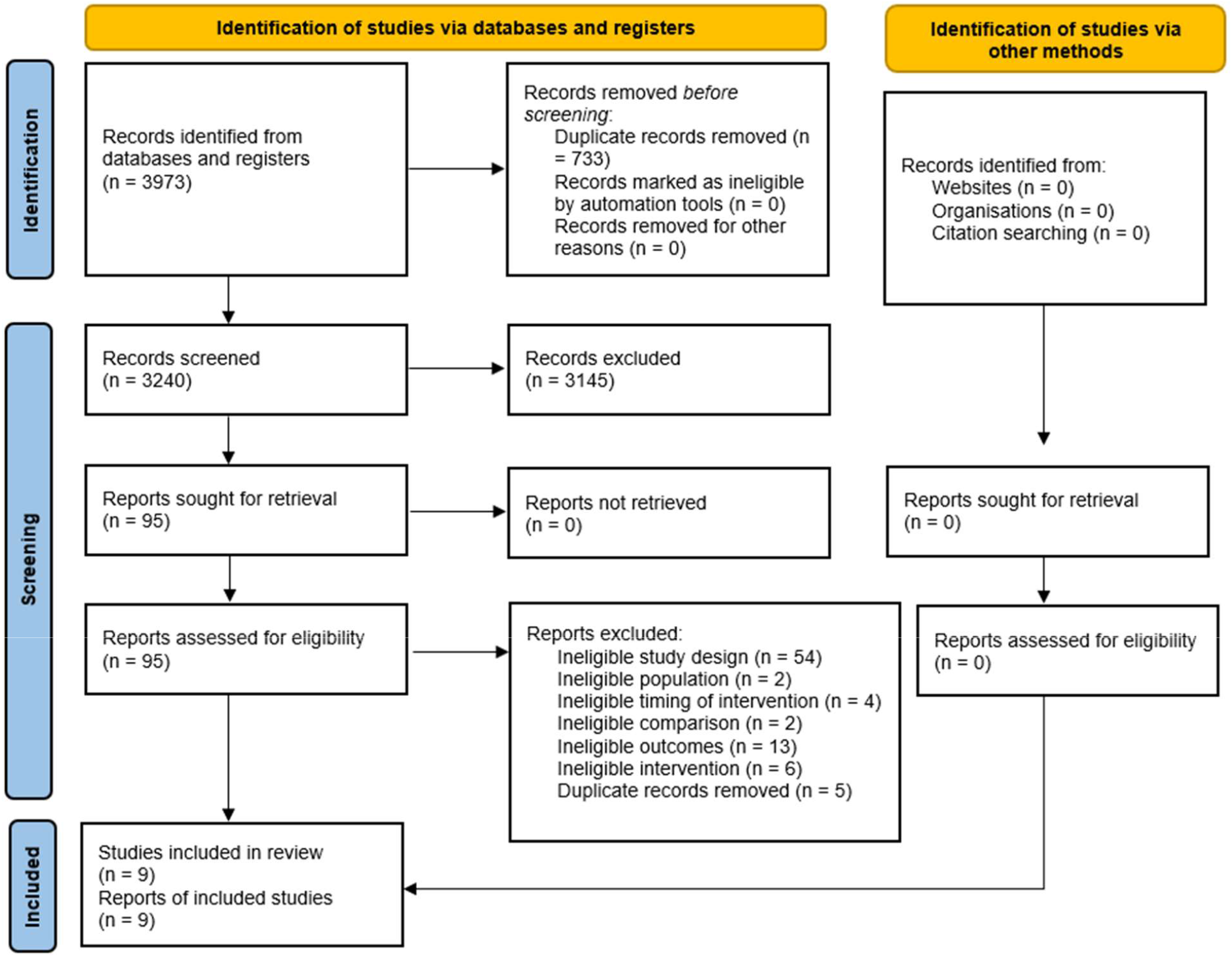
Literature search flow and study selection diagram

The main characteristics of included studies are summarized in Table 1. A total of 1273 subjects were included from 9 RCTs. Three studies were conducted in the United States^18–20^, 2 studies in Iran^21, 22^, 1 study in Bangladesh,^23^ 1 study in India,^24^ 1 unpublished study in Brazil (DIUPRE), and 1 in the UK.^25^ Patients in the intervention group received diuretics for a period of 5 to 7 days. All studies assigned their intervention group to furosemide 20-40 mg daily except for Viteri et al.^19^ who assigned intervention group patients to torsemide 20 mg daily. Matthews et al.^25^, Viteri et al.^19^, Perdigao et al.^20^, and the DIUPRE study assigned their control groups to placebo. Three studies assigned their control groups to alternative antihypertensive medications. The control groups in Veena et al.^18^ and Bozorgan et al.^22^ were assigned to nifedipine 10 mg every 8 hours and the control group in Siamansoori et al.^21^ was assigned to methyldopa 250 mg three times daily. The control groups in Ascarelli et al.^18^ and Dabaghi et al.^23^ were assigned to no medication. The mean number of study participants was 115. The trials with the largest number of subjects had 384^20^ and 264^18^ subjects, respectively. The study with the least number of participants had 19 subjects.^25^

**Table 1.** Main characteristics of included studies

| Year, author | Country | Included HDP | Important exclusion criteria | Total (n) | Intervention (n) | Comparison (n) | Primary outcome | Secondary outcomes |
| --- | --- | --- | --- | --- | --- | --- | --- | --- |
| Matthews et al<sup>25</sup> (1997) | UK | Preeclampsia | Diabetes mellitus, renal or hepatic impairment, contraindication to diuretic | 19 | Furosemide 40 mg orally every 24 hours for 7 days (10) | Placebo orally every 24 hours for 7 days (8) | Mean and maximum BP | Postpartum antihypertensive usage<br>Length of hospital stay |
| Ascarelli et al<sup>18</sup> (2005) | USA | Chronic HTN, gestational HTN, preeclampsia | Less than 20 weeks of gestation, hypokalemia ($K < 3.0$ mEq/L), already taking diuretics or potassium supplementation, hemodynamic instability | 264 | Furosemide 20 mg orally every 24 hours for 5 days (132) | No medication (132) | Systolic and diastolic BP | Length of hospital stay<br>Postpartum antihypertensive usage |
| Veena et al<sup>24</sup> (2017) | India | Severe preeclampsia | Hemodynamic instability, hypokalemia ($K < 3$ mEq/L), already on potassium supplementation and diuretics, expulsion of the fetus at $< 20$ weeks | 100 | Furosemide 20 mg orally every 24 hours with nifedipine 10 mg orally every 8 hours until not needed (50) | Nifedipine 10 mg every 8 hours until not needed (50) | Reduction in mean, systolic, and diastolic BP | Length of hospital stay<br>Postpartum antihypertensive usage<br>Antihypertensive requirement at discharge |
| Viteri et al<sup>19</sup> (2018) | USA | Preeclampsia with or without severe features, superimposed preeclampsia with or without severe features | Oliguria, heart failure, hypokalemia (serum potassium below 3 mEq/L), diuretic use within the past 24 hours, hypersensitivity to torsemide or sulfonylureas, and pulmonary edema | 118 | Torsemide 20 mg orally every 24 hours for 5 days (59) | Placebo orally every 24 hours for 5 days (59) | BP $> 150/100$ by day 5 or hospital discharge | Persistent HTN at discharge 7–10 days and 6 weeks after delivery<br>Postpartum antihypertensive usage,<br>Length of hospital stay<br>Hospital readmission for HTN |
|  |  |  |  |  |  |  |  | Adverse events to diuretics |
| Dabaghi et al<sup>23</sup> (2019) | Bangladesh | Severe preeclampsia HELLP syndrome | Gestational age under 20 weeks, hypokalemia ($k < 3.0$ mEq/L) on admission, history of diuretics or potassium supplements use, any hemodynamic instability surrounding the events of delivery | 90 | Furosemide 20 mg and potassium supplementation every 24 hours for 5 days (45) | No medication (45) | Mean systolic BP | Postpartum antihypertensive usage<br>Length of hospital stay<br>Eclampsia |
| Siamansoori et al<sup>21</sup> (2020) | Iran | Preeclampsia | Underlying cardiac disease, complicated pregnancies including gestational diabetes mellitus, renal disease, hypersensitivity to furosemide or methyldopa, major adverse drug complications | 100 | Furosemide 20 mg twice daily (50) | Methyldopa 250 mg three times daily (50) | Systolic and diastolic BP at 6 hours, 48 hours and one week | Diuresis at 24 and 48 hours post intervention |
| Perdigao et al<sup>20</sup> (2021) | USA | All forms of HDP: gestational HTN, preeclampsia with or without severe features, preeclampsia superimposed on chronic HTN with or without severe features | Underlying cardiac disease, rheumatologic disease, advanced diabetes, elevated creatinine ($>1.2$ mg/dL), significant hypokalemia ($K < 3$ mEq/L), allergy to furosemide, or those who received diuretics before randomization | 384 | Furosemide 20 mg orally every 24 hours for 5 days (192) | Placebo orally every 24 hours for 5 days (192) | Persistent HTN 7 days postpartum (defined as at least 2 consecutive BP readings over 48 hours of systolic BP $\geq 140$ mm Hg and diastolic BP $\geq 90$ mm Hg) and the number of days required to achieve resolution of HTN (defined as at least 2 consecutive BP readings over 48 hours of systolic BP $< 140$ mm Hg) | Percentage of postpartum BP in the severe range (systolic BP $\geq 160$ mm Hg or diastolic BP $\geq 110$ mm Hg)<br>Postpartum antihypertensive usage<br>Hospital readmissions or ER visits for HTN<br>Length of hospital stay<br>HTN complications |
|  |  |  |  |  |  |  | and diastolic BP<br><90 mm Hg) |  |
| Bozorgan, et al<sup>22</sup> (2022) | Iran | Preeclampsia | Chronic HTN, BP less than 150/100 mmHg, administration of diuretics, renal disease, diabetes, hemodynamic instability, potassium level lower than 3 mEq/L, contraindication to furosemide, hematocrit more than 37% | 116 | Nifedipine 10 mg every 8 hours and furosemide 20 mg daily for 5 days (58) | Nifedipine 10 mg every 8 hours for 5 days (58) | Systolic and diastolic BP at day 2-5 | Postpartum antihypertensive usage |
| DIUPRE (2024) | Brazil | Preeclampsia | Chronic HTN, blood pressure <140/90 mmHg, diuretic use, renal impairment, diabetes, sickle cell disease, rheumatologic disease, hemodynamic instability, potassium less than 3 mEq/L, contraindication for furosemide use | 120 | Furosemide 40 mg orally every 24 hours for 5 days (60) | Placebo orally every 24 hours for 5 days (60) | Mean blood pressure from 24 hours after delivery to first 15 days of delivery | Postpartum antihypertensive usage<br>Time to blood pressure control<br>Daily urine output<br>Length of hospital stay<br>Frequency of adverse events<br>Frequency of maternal complications |

BP: blood pressure; HDP: hypertensive disorders of pregnancy; HTN: hypertension; MAP: mean arterial pressure; ER: emergency
room; HELLP: hemolysis, elevated liver enzyme, low platelet count

Primary outcomes varied between studies and included mean systolic and diastolic BPs, reduction in mean BPs, and persistent HTN at various time points up to 10 days post-hospital discharge. Observation periods varied among studies and lasted up to 6 weeks post-delivery and post-hospitalization.

### Primary Outcome

The primary outcome, persistent HTN at 0-10 days post-delivery, was available in 7 studies.^18–25^ Persistent HTN was significantly less frequent in the intervention group for the two most recent studies,^20, 22^ including the largest study in our analysis.^20^ However, in our random effects model there was no significant difference in persistent HTN among individuals who received postpartum diuretics compared with controls (odds ratio [OR]: 0.70; 95% confidence interval [CI]: 0.46-1.05), shown in Figure 2. Study heterogeneity was moderate for the primary outcome (I^2 of 40%, tau ^2 0.1208, p=0.12).

**Figure 2.**
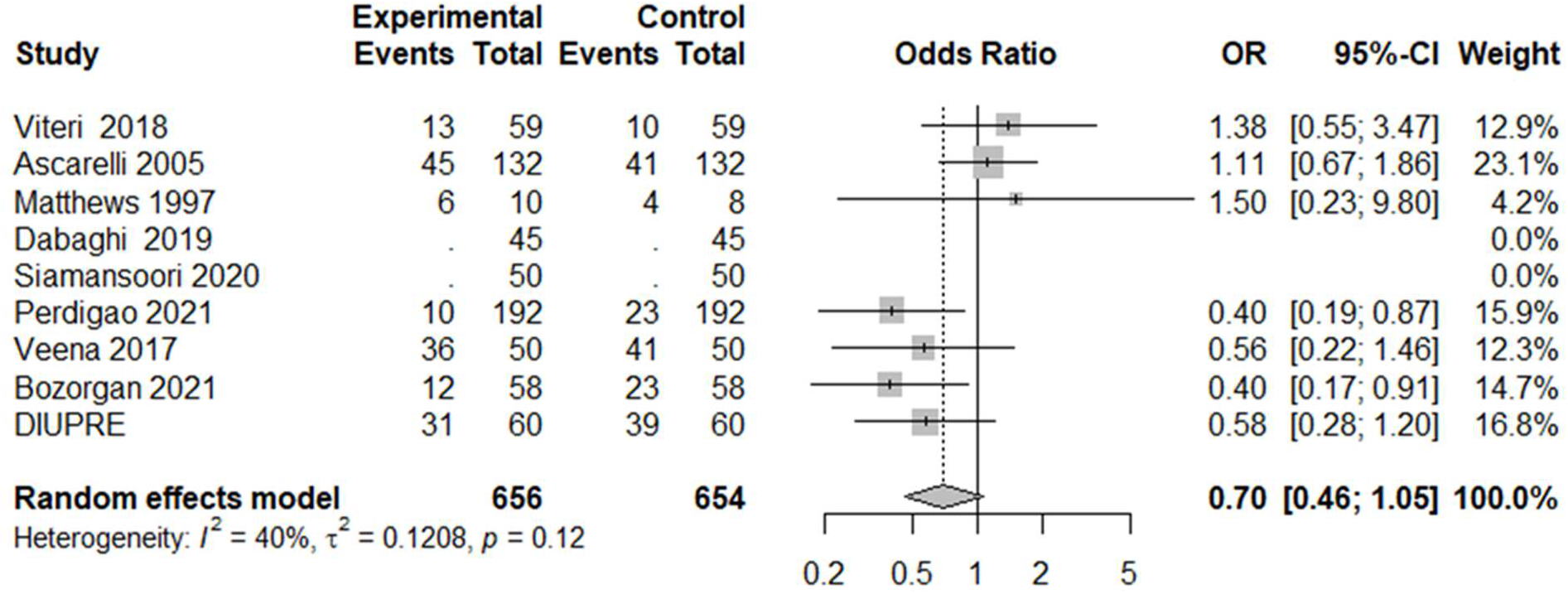
Persistent hypertension at 0-10 days postpartum among individuals with hypertensive disorders of pregnancy treated with diuretic versus control OR: odds ratio; CI: confidence interval

## Secondary Outcomes

### Systolic and diastolic blood pressure in the postpartum period

The systolic and diastolic BP measurements were available for 3 studies which encompassed 23% of patients included in the analysis (n=306).^21–23^ BP measurements were monitored for 3 to 7 days postpartum. Postpartum diuretic use was associated with lower systolic BP (square mean difference [SMD]: -0.36; 95% CI: -0.72; -0.01) (Figure 3) without a difference in diastolic BP (SMD: 0.01; 95% CI: -0.22; 0.23) (Figure 4) compared with controls. The lower systolic BP effect was largest in the Bozorgan et al. study.

**Figure 3.**
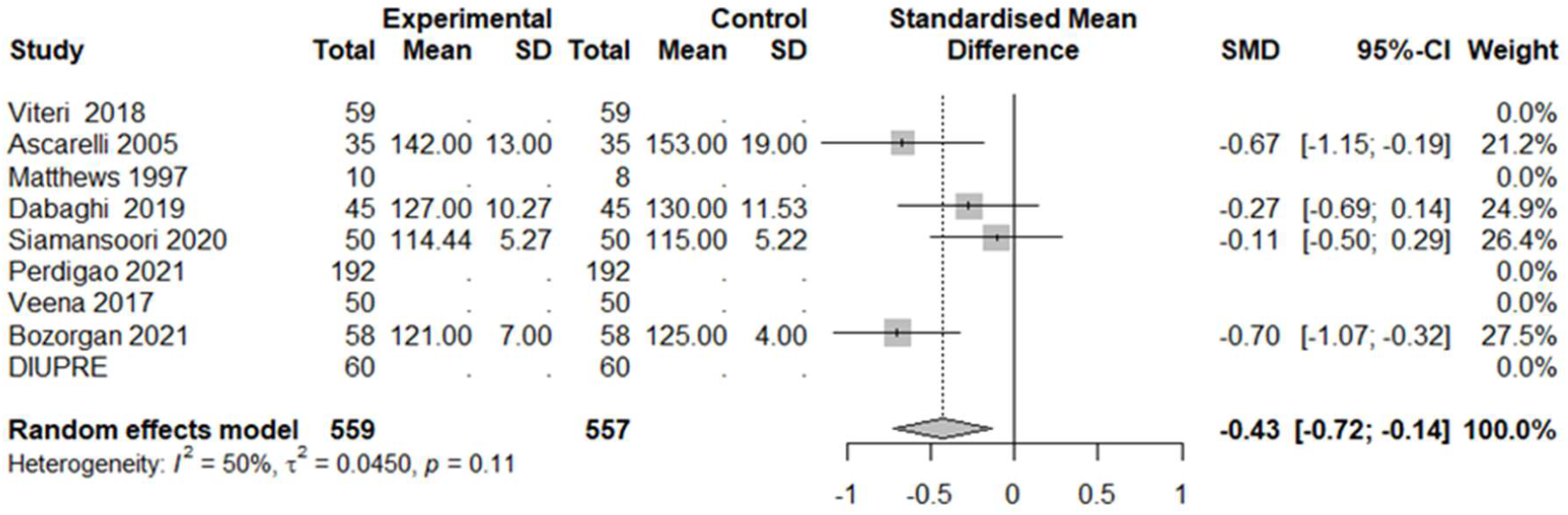
Mean difference in systolic blood pressure among individuals with hypertensive disorders of pregnancy with postpartum diuretic treatment compared with control SD: standard deviation; SMD: standardized mean difference; CI: confidence interval

**Figure 4.**
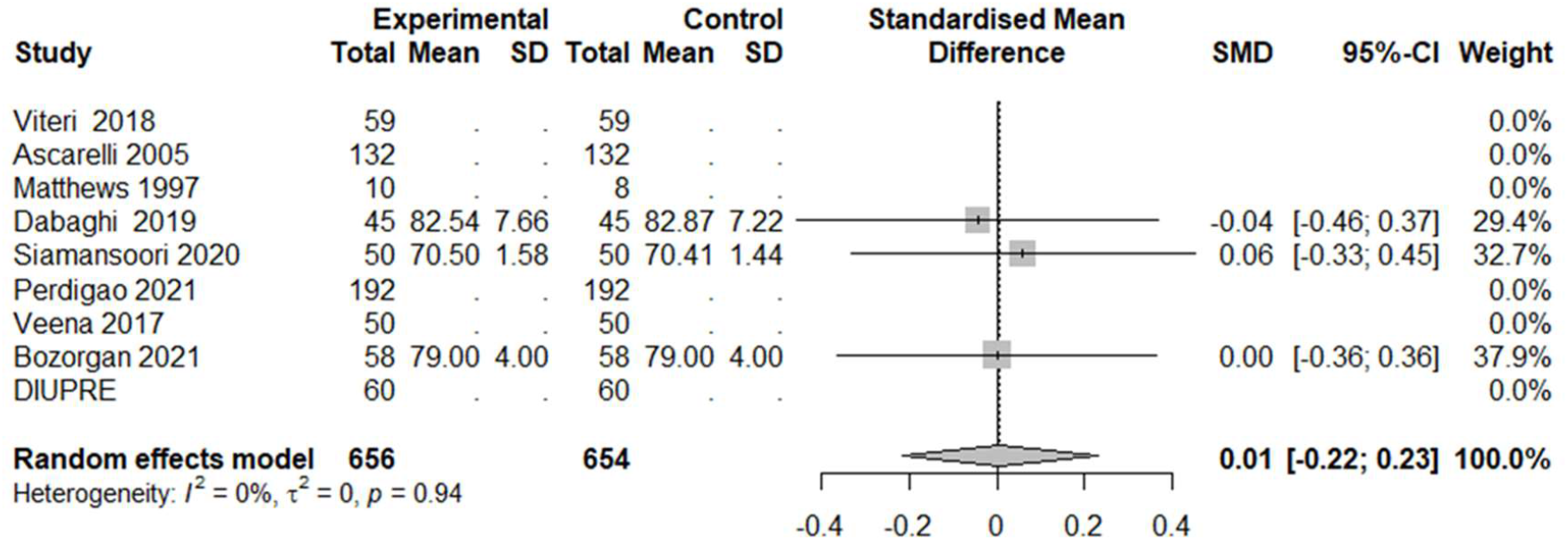
Mean difference in diastolic blood pressure among individuals with hypertensive disorders of pregnancy with postpartum diuretic treatment compared with control SD: standard deviation; SMD: standardized mean difference; CI: confidence interval

### Antihypertensive use postpartum

Additional antihypertensive medication use during an observation period up to 10 days was reported in 8 studies (Figure 5). Although Veena et al. and Bozorgan et al. reported significantly lower use of additional antihypertensive drugs in the intervention group compared with the control group, there was no significant difference in the remaining studies.^22, 24^ This effect was largest in the Bozorgan et al. study (OR: 0.23; 95% CI: 0.08-0.67). Pooling data from all 8 studies, there was no significant difference in postpartum antihypertensive use in patients who received diuretics compared with controls (OR: 0.66; 95% CI: 0.42-1.05; p=0.08). Study heterogeneity was moderate with I^2 of 34%.

**Figure 5.**
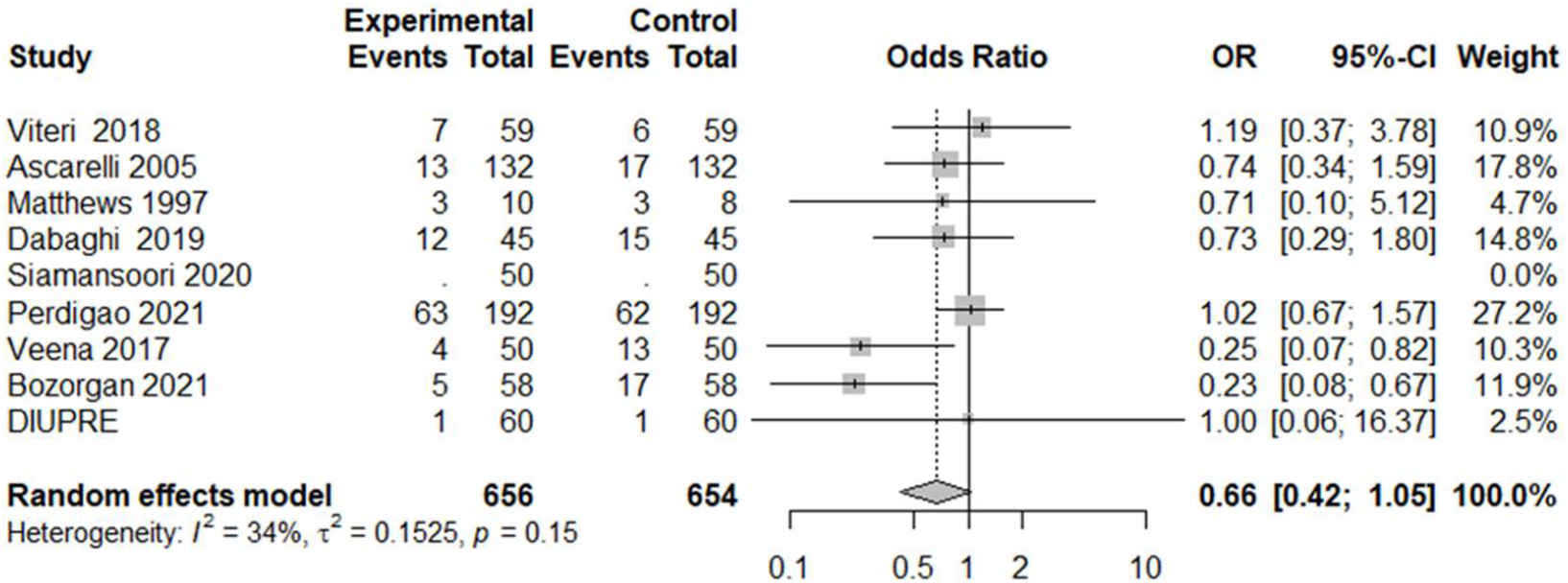
Use of additional antihypertensive medication among individuals with hypertensive disorders of pregnancy treated with diuretic compared with control OR: odds ratio; CI: confidence interval

### Length of hospital stay

Although 6 studies commented on length of hospital stay, there was significant heterogeneity in these reports which precluded the ability to calculate a pooled effect size for this secondary outcome. In brief, three studies reported postpartum hospital length of stay^19, 20, 24^ while three others reported total hospital length of stay.^18, 23, 25^ No studies reported a significant difference in these outcomes between treatment groups.

### Postpartum maternal complications

There was limited reporting of our additional secondary outcome variables of interest including hospital readmission, emergency department visits, heart failure, b-type natriuretic peptide levels, and lactation outcomes. Therefore, analyses were not performed on these outcomes.

Three studies did report outcomes related to emergency department visits or hospital readmissions without any significant differences between treatment groups, however their variations in outcome definitions and the inclusion of a CI or standard error in only one of these studies precluded the calculation of pooled estimates.^19, 20, 25^

### Risk of Bias Assessment Results

Detailed study quality assessments are provided in Table 2. Five studies received a low risk of bias.^19, 20, 24, 25^ Two studies received some concerns of risk of bias around reporting of missing outcome data and bias around measurement of the outcomes.^18, 21^ Lastly, two studies received an overall high risk of bias for significant concerns with multiple domains including the randomization process, intervention assignment, missing outcome data, and outcome measurement.^22, 23^

**Table 2.** Risk of bias assessments of included studies

| Studies | Risk of bias domains |  |  |  |  | Overall risk of bias |
| --- | --- | --- | --- | --- | --- | --- |
|  | Randomization process | Assignment to intervention | Missing outcome data | Measurement of the outcome | Selection of the reported result |  |
| Matthews et al<sup>25</sup> (1997) | Low | Low | Low | Low | Low | Low |
| Ascarelli et al<sup>18</sup> (2005) | Low | Low | Some concerns | Some concerns | Low | Some concerns |
| Veena et al<sup>24</sup> (2017) | Low | Low | Low | Low | Low | Low |
| Viteri et al<sup>19</sup> (2018) | Low | Low | Low | Low | Low | Low |
| Siamansoori et al<sup>21</sup> (2019) | Low | Low | Some concerns | Low | Low | Some concerns |
| Dabaghi et al<sup>23</sup> (2019) | Low | Some concerns | Some concerns | Some concerns | Low | High |
| Bozorgan et al<sup>22</sup> (2021) | Some concerns | Some concerns | Low | Low | Low | High |
| Perdigao et al<sup>20</sup> (2021) | Low | Low | Low | Low | Low | Low |
| DIUPRE (2024) | Low | Low | Low | Low | Low | Low |

## DISCUSSION

In this meta-analysis, we assessed the role of postpartum diuretics after HDP on improving postpartum HTN and BP. We found that postpartum diuretic use was not associated with reduced postpartum hypertension but was associated with reduced systolic BP compared with controls without a statistically significant difference diastolic BP, or use of additional antihypertensive medications. However, the available data to assess the impact of postpartum diuretics in individuals with HDP remains limited. The number of available studies was relatively small, with some studies including only a small number of patients and some with concerns about risk of bias. Despite these limitations, there was a non-significant trend of reduced persistent postpartum HTN with an OR of 0.70 and of reduced use of antihypertensive medications with an OR of 0.66. While these findings did not reach statistical significance, they indicate potentially impactful trends. A well-conducted, sufficiently powered RCT could provide more definitive data on the potential benefit of postpartum diuretics in this population.

A recent systematic review and meta-analysis on the use of loop diuretics in the context of HDP similarly found no reduction in persistent HTN at 1 or 6 weeks postpartum and no reduction in the need for additional antihypertensive medications at discharge.^7^ Our meta-analysis expands on these findings with the inclusion of three additional studies. We assessed 9 trials comparing loop diuretics to controls, including the unpublished results of one completed registered clinical trial. Most of the included trials were individually limited by smaller sample size and short-term follow-up.

Our finding of reduced systolic BP is novel and although the effect size is small, improved BP control in the immediate short-term postpartum period may confer durable clinical benefit. Pharmaceutical intervention in the immediate postpartum period has been shown to result in improved BP control sustained up to 3-4 years postpartum^26, 27^ and be associated with more favorable left atrial and left ventricular remodeling, suggesting that prompt postnatal BP control may reverse the adverse remodeling known to result from HDP.^28^ Nonetheless, the lack of a diastolic BP benefit is relevant in this patient population. The association between HDP and later-life cardiovascular disease is partially mediated through the development of chronic HTN, with up to 37% of individuals with a HDP developing chronic HTN at 2-7 years after delivery.^29–31^ In a study of BP trajectories in the first year postpartum following a HDP, the majority of those with persistent HTN had isolated diastolic HTN, which may suggest that targeting an improvement in postpartum diastolic BP is of particular importance following an HDP.^32^

Efficacy of postpartum diuretics may vary by severity of HDP. Perdigao et al. found that reduction in persistent HTN at 7 days postpartum was more significant when results were stratified to only individuals with non-severe HDP. Interestingly, there was no difference in persistent HTN in the severe HDP group (systolic BP ≥160 mm Hg or diastolic BP ≥110 mm Hg).^20^ In contrast, Ascarelli et al. found that reduction in persistent HTN 2 days postpartum was more significant in patients with severe preeclampsia only.^18^ There was heterogeneity among the studies in their inclusion of only severe versus both severe and non-severe HDP, and future studies should give particular attention to or perform subgroup analyses for these subgroups of HDP severity to define which group may particularly benefit from postpartum diuretics. There was also variation in the spectrum of HDP subtypes which were included in the studies. Importantly, some studies included chronic HTN which may be less diuretic responsive with regards to BP recovery and thus attenuate the measured benefit by the total studied population of individuals with HDP.

Ongoing RCTs will hopefully contribute to narrowing knowledge gaps about the benefits of postpartum diuretics among individuals with HDP. The largest randomized placebo-controlled trial of postpartum diuretics in individuals with HDP will include an estimated 612 participants and will compare hydrochlorothiazide 50 mg daily to placebo for 14 days after delivery (NCT03298802). Primary outcomes include need for additional antihypertensive drug and rate of readmission or triage visits 1-6 weeks postpartum. As our meta-analysis included only 1273 subjects, the results of this trial would expand the number of subjects to more definitively assess the utility of postpartum diuretics in HDP, however the effect on blood pressure may be different with thiazide diuretics compared with loop diuretics.

There may also be a role for postpartum diuretics for prevention of HDP. The recently published Lasix for the Prevention of De Novo Postpartum Hypertension (LAPP) randomized placebo-controlled trial extends our understanding of the preventive role of postpartum diuretics by comparing primary prevention furosemide to placebo among 82 individuals at high risk for de novo postpartum HTN. The LAPP study found no difference in mean arterial pressure 24 hours prior to discharge or in the development of de novo postpartum HTN between the two groups.^33^ As 42% of individuals with postpartum hypertension have preceding antepartum normotension, the identification of individuals at high risk to develop postpartum HTN and strategies to mitigate associated volume overload are needed.^34^

### Limitations

The RCTs available for inclusion in our meta-analysis resulted in a modest sample size. Two of the studies, accounting for 16% of the patients included in the meta-analysis, were felt to have high risk of bias. There was heterogeneity in the control groups, with only four of the studies comparing diuretic to placebo and the remaining studies comparing diuretic to alternative antihypertensive or no medication, which could bias results towards the null hypothesis. Systolic and diastolic BP data were available from only three studies for inclusion in our analysis.

Further, due to the lack of standardized guidance for postpartum BP monitoring, there was notable variation in the timing of study outcome assessments. Some studies reported this data at postpartum day 0-5, others at days 5-10, and others at 30 days. Consequently, study outcome heterogeneity was moderate per our random effect size analysis specifically regarding persistent postpartum HTN. Our analysis may have been affected by this heterogeneity in BP recording postpartum.

## Conclusions

The data on the benefit of postpartum oral diuretics for the prevention of postpartum HTN among women with HDP remains inconclusive. Diuretic administration reduces systolic BP and may reduce the risk of HTN up to 10 days postpartum for some clinical subgroups. However, due to the low certainty of evidence, uniform postpartum use of diuretics among women with HDP cannot be recommended. Use of postpartum diuretics guided by thoughtful assessment of clinical volume status remains reasonable.

## Data Availability

All previously published data utilized in this systematic review and meta-analysis were obtained from publicly available sources. Unpublished data from the DIUPRE study were obtained from trial investigators. Data is available from the corresponding author upon reasonable request.

## ACKNOWLEDGEMENTS

None.

## ABBREVIATIONS

HDP: hypertensive disorder of pregnancy
HTN: hypertension
BP: blood pressure
RCT: randomized controlled trials

---

**Supplement Table 1.**
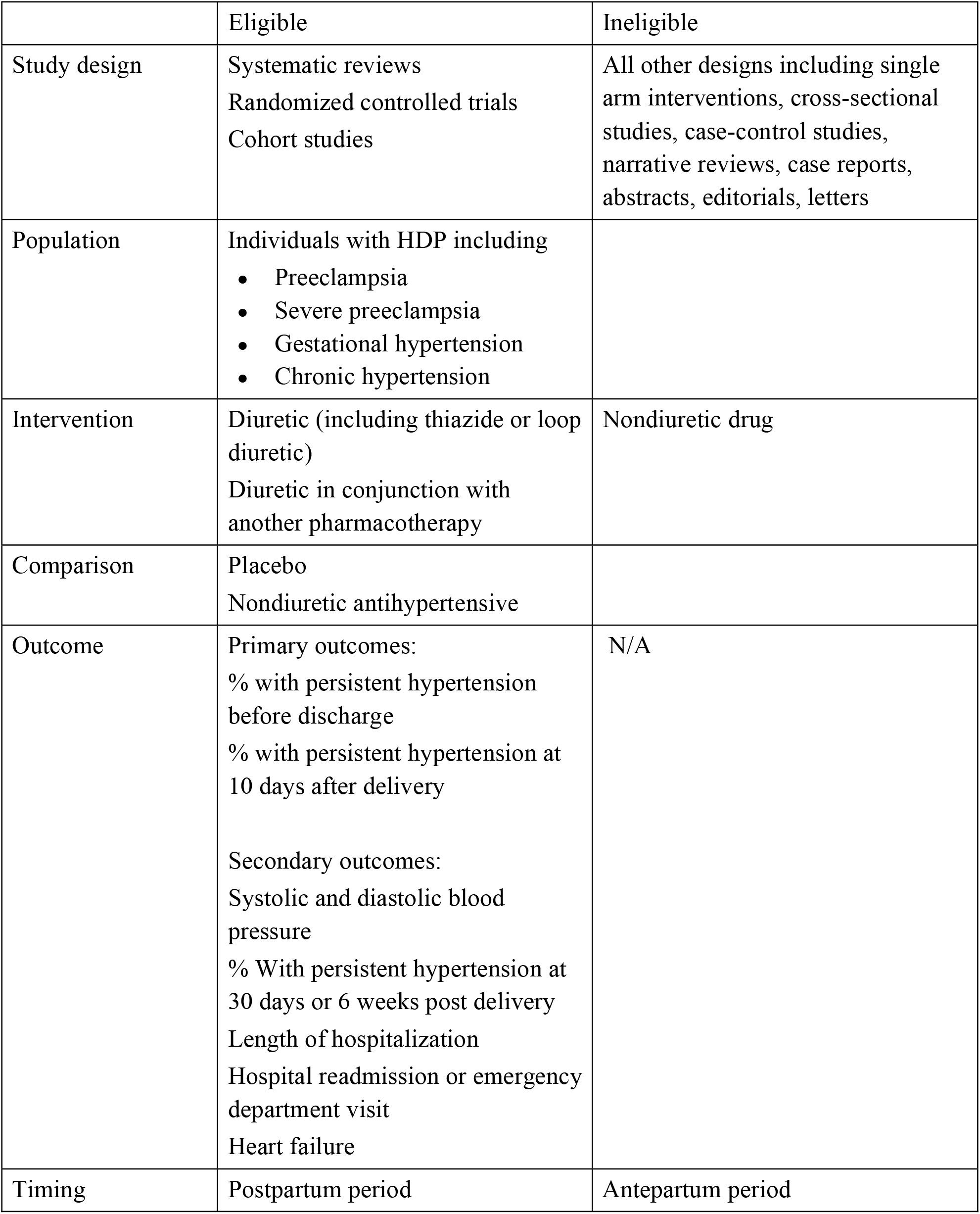
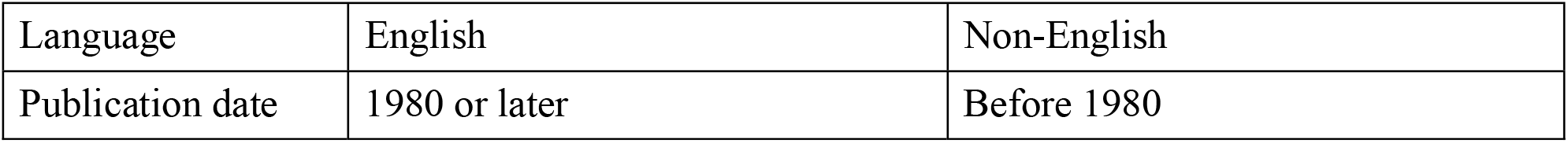
Study selection criteria based on study design, population, interventions, comparators, outcomes, timing

## Search strategies

All searches performed on May 11, 2021 and reran on June 29, 2022 <u>Ovid Medline 1</u>

Embase.com 2 Web of Science Core Collection 5

Cochrane Central Register of Controlled Trials (CENTRAL) 8

Clinicaltrials.gov 10

## Ovid Medline

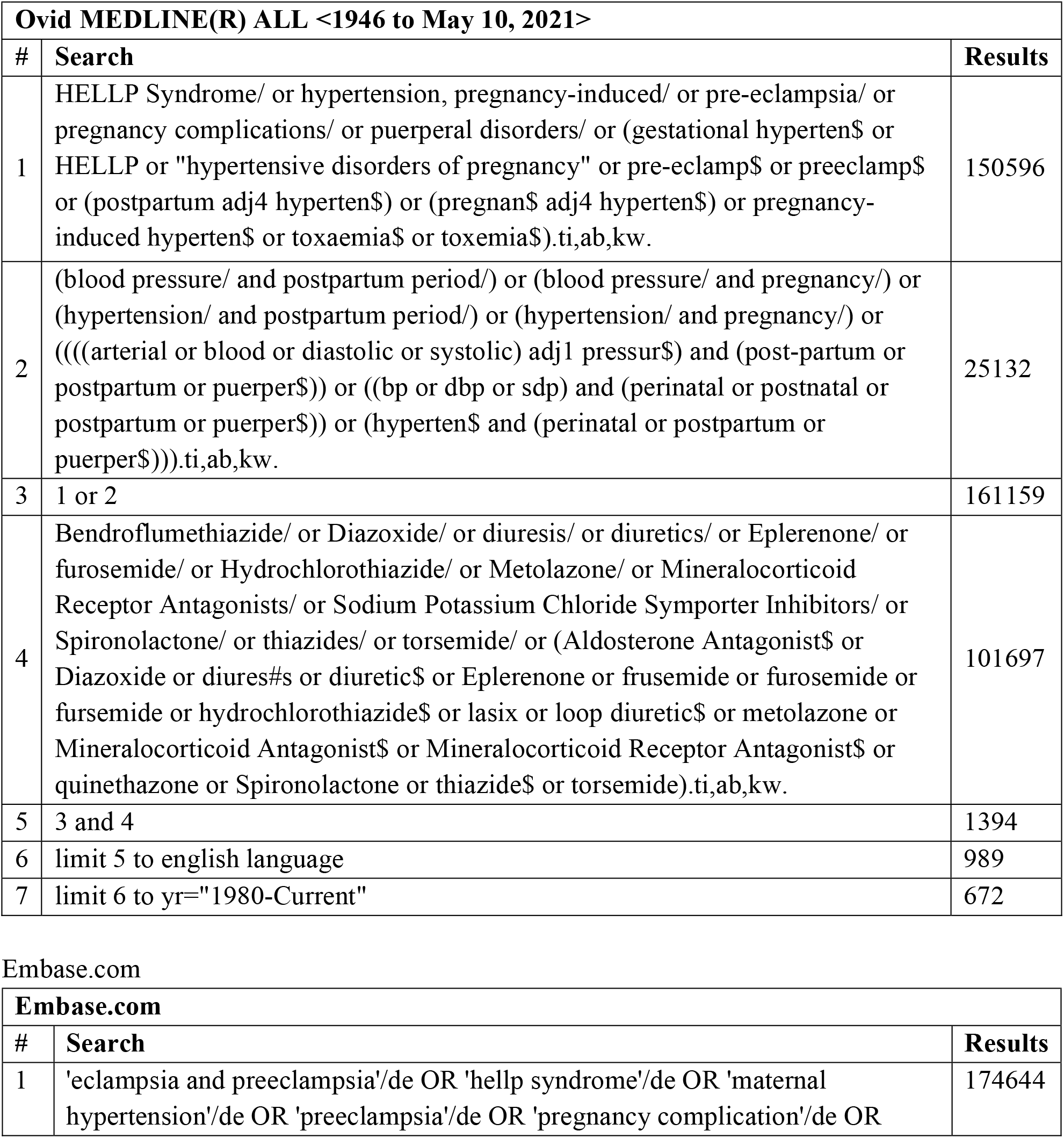
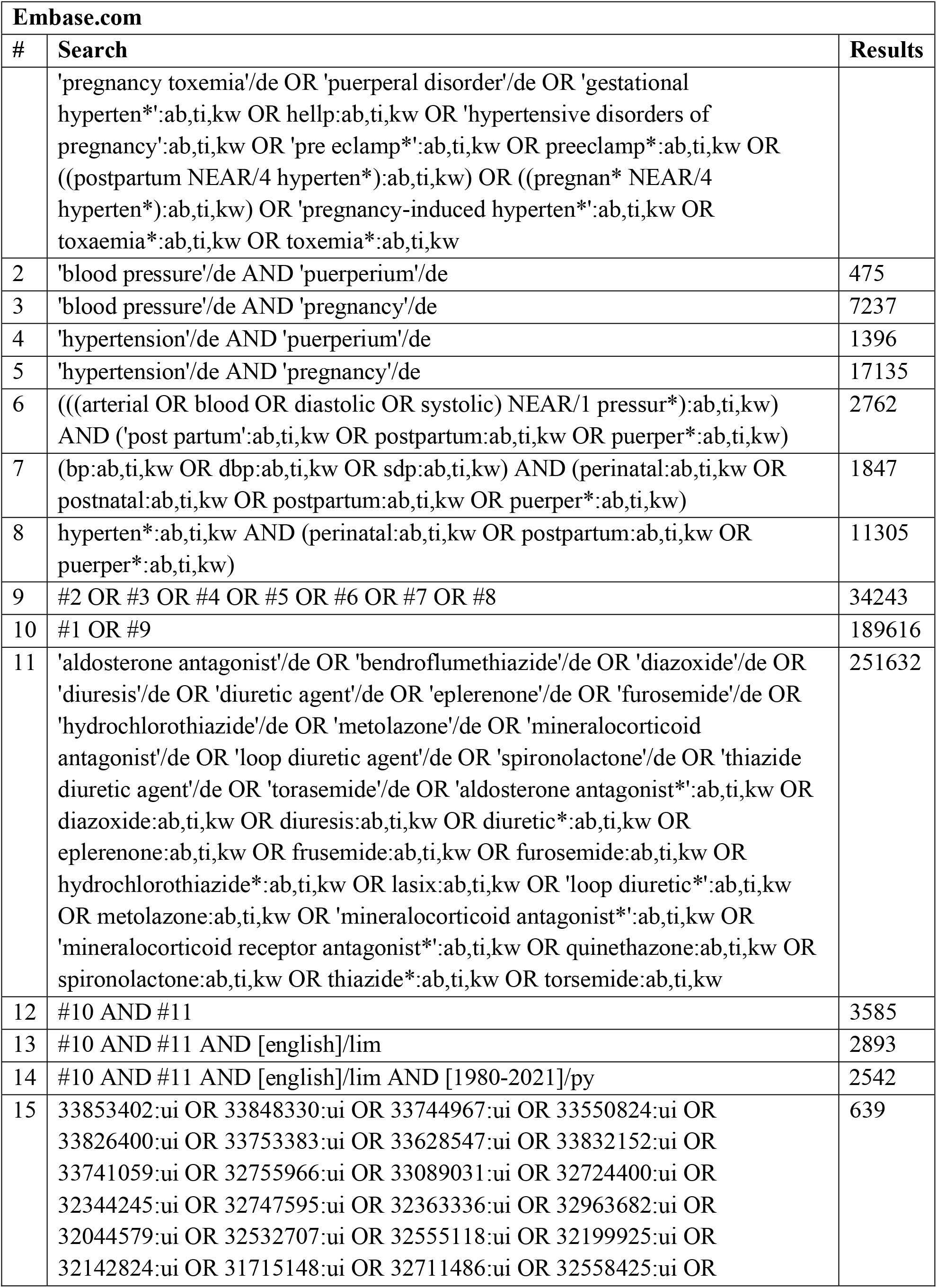
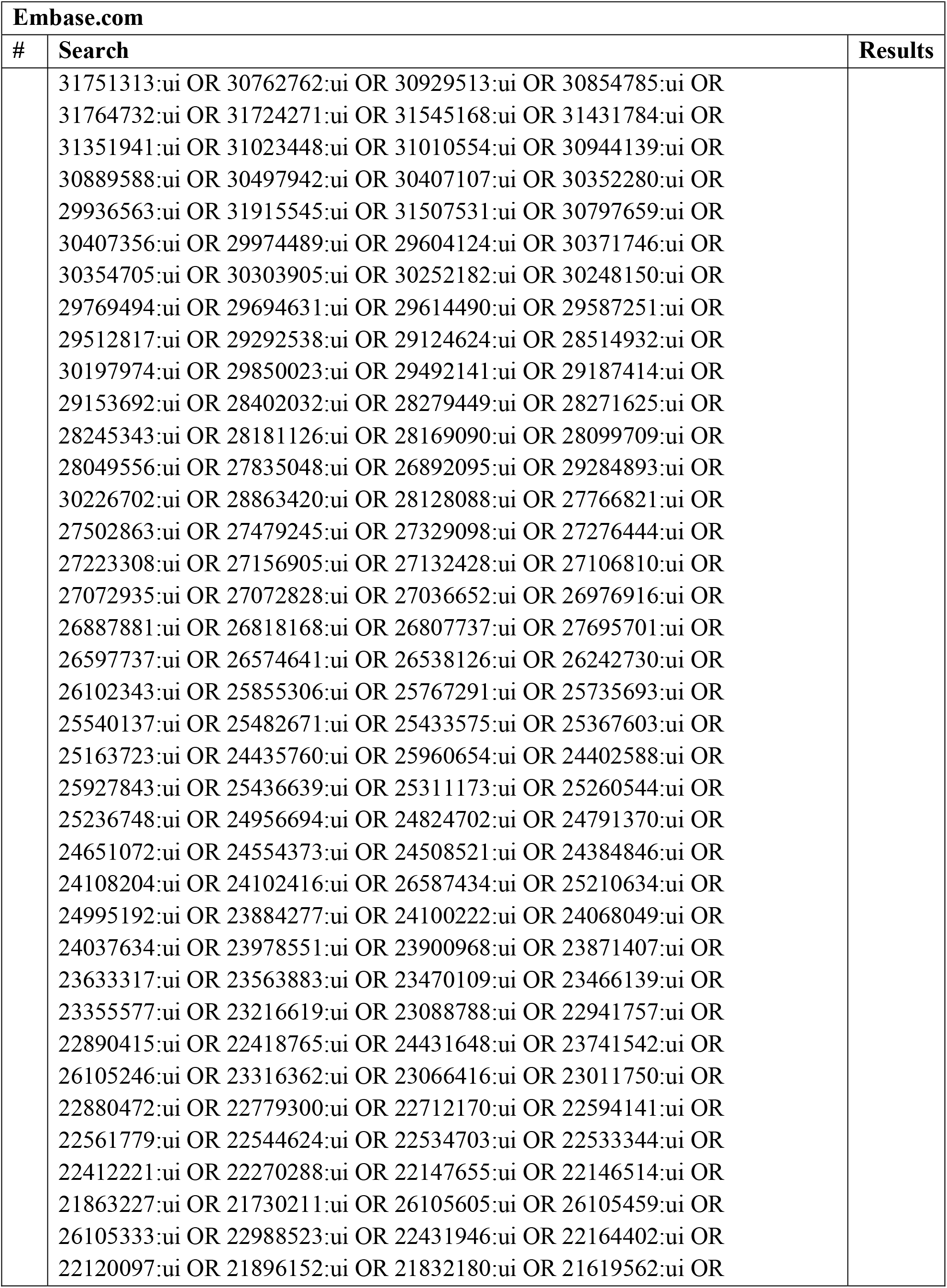
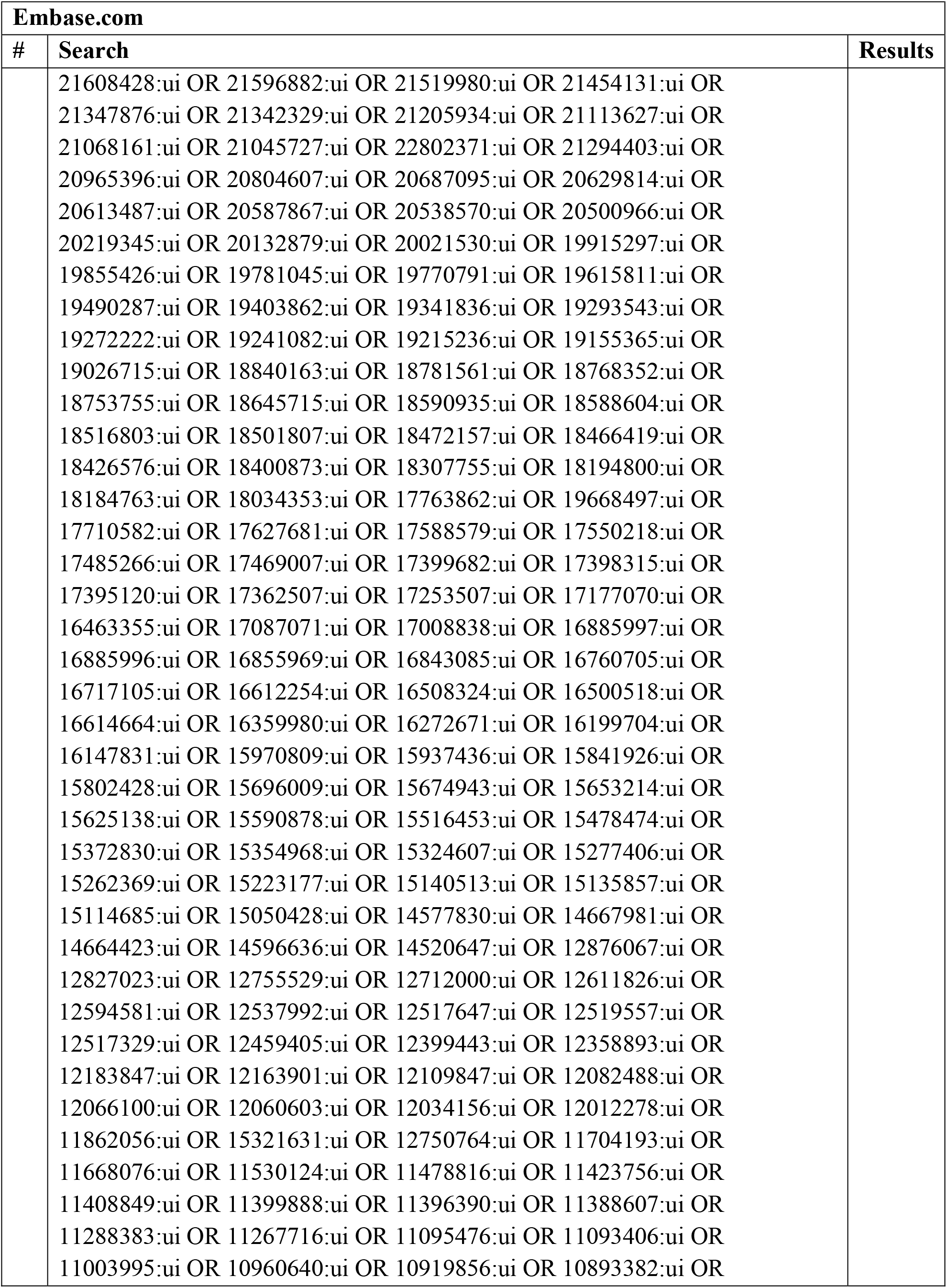
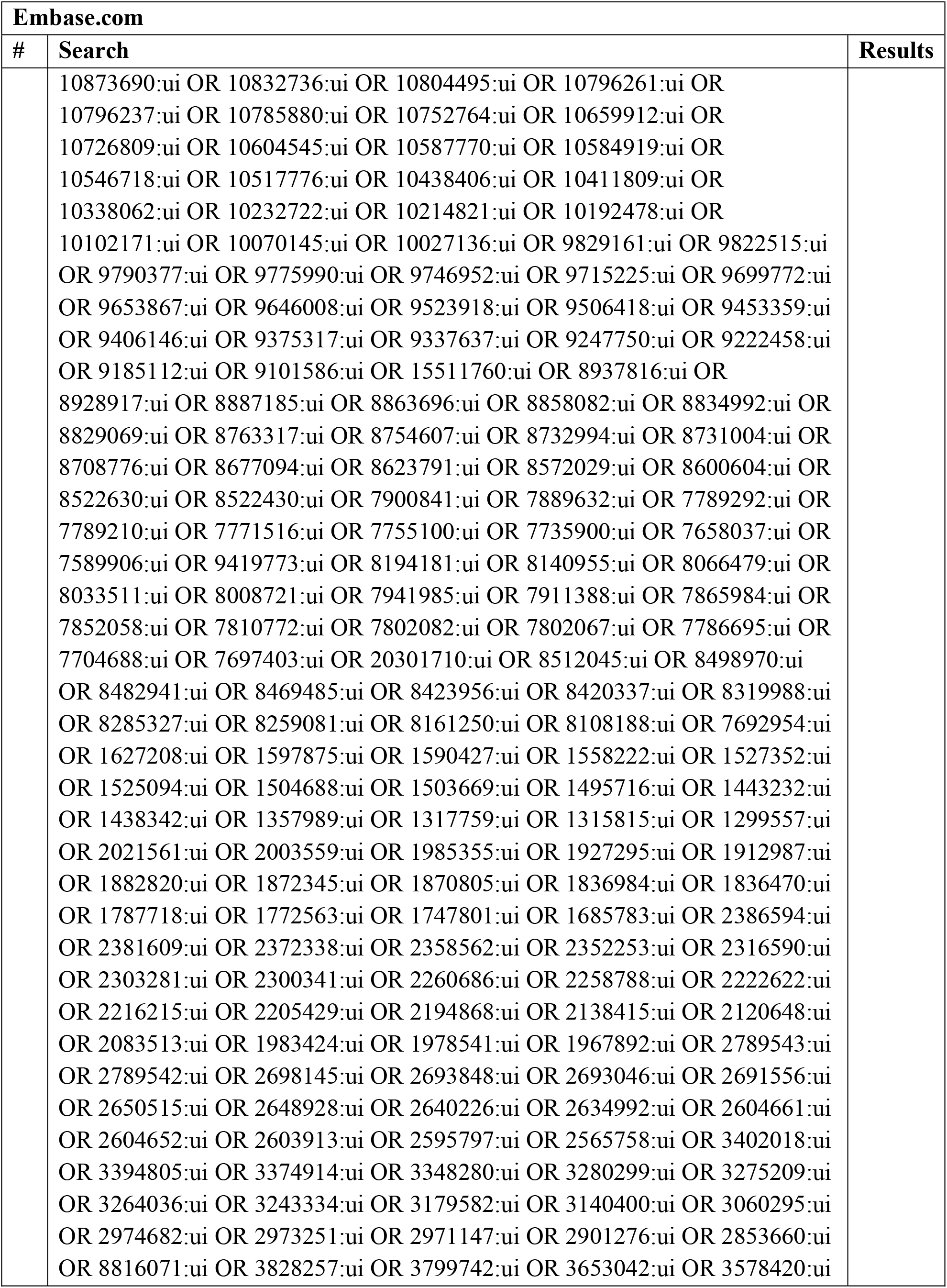
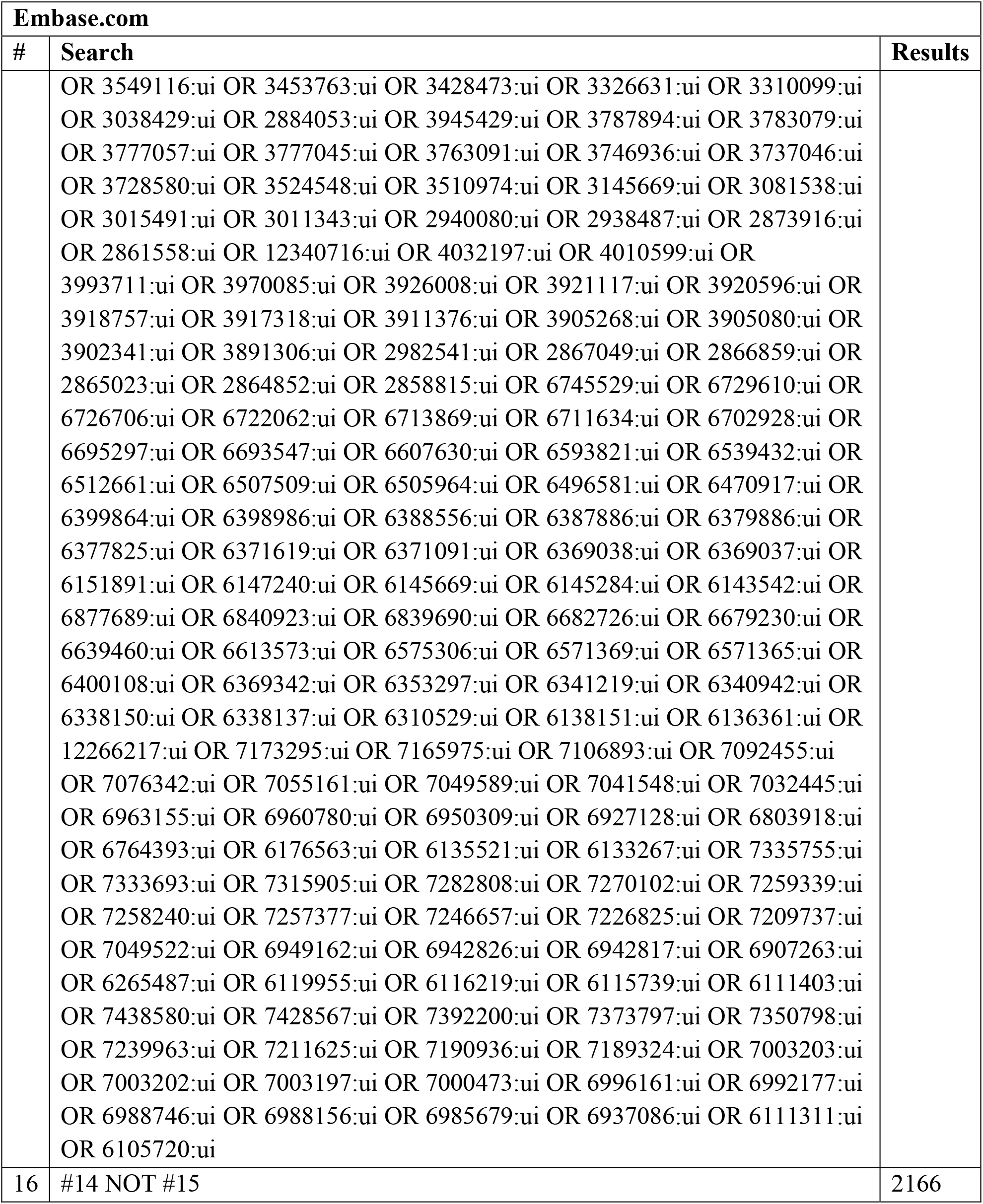

## Web of Science Core Collection

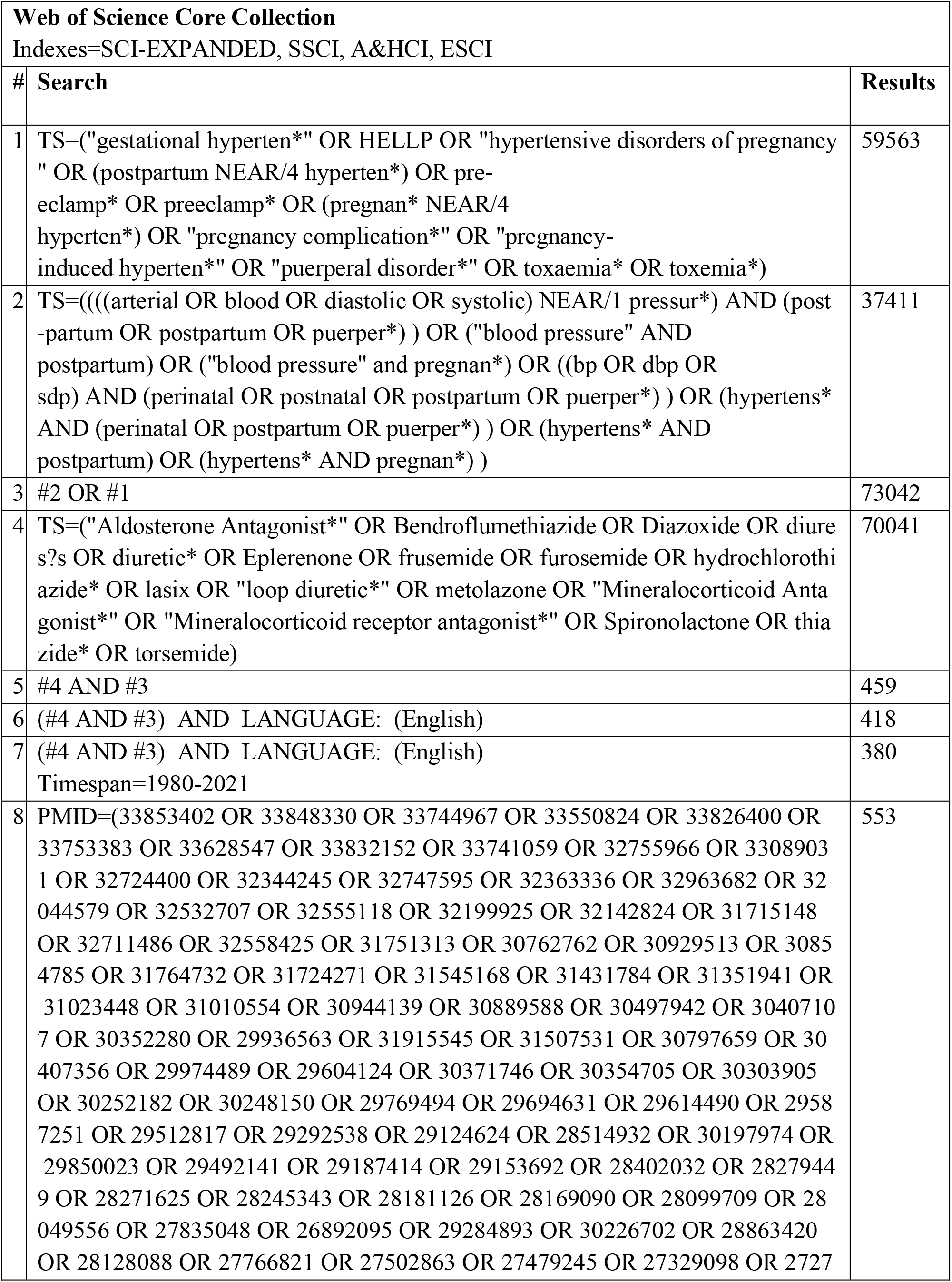
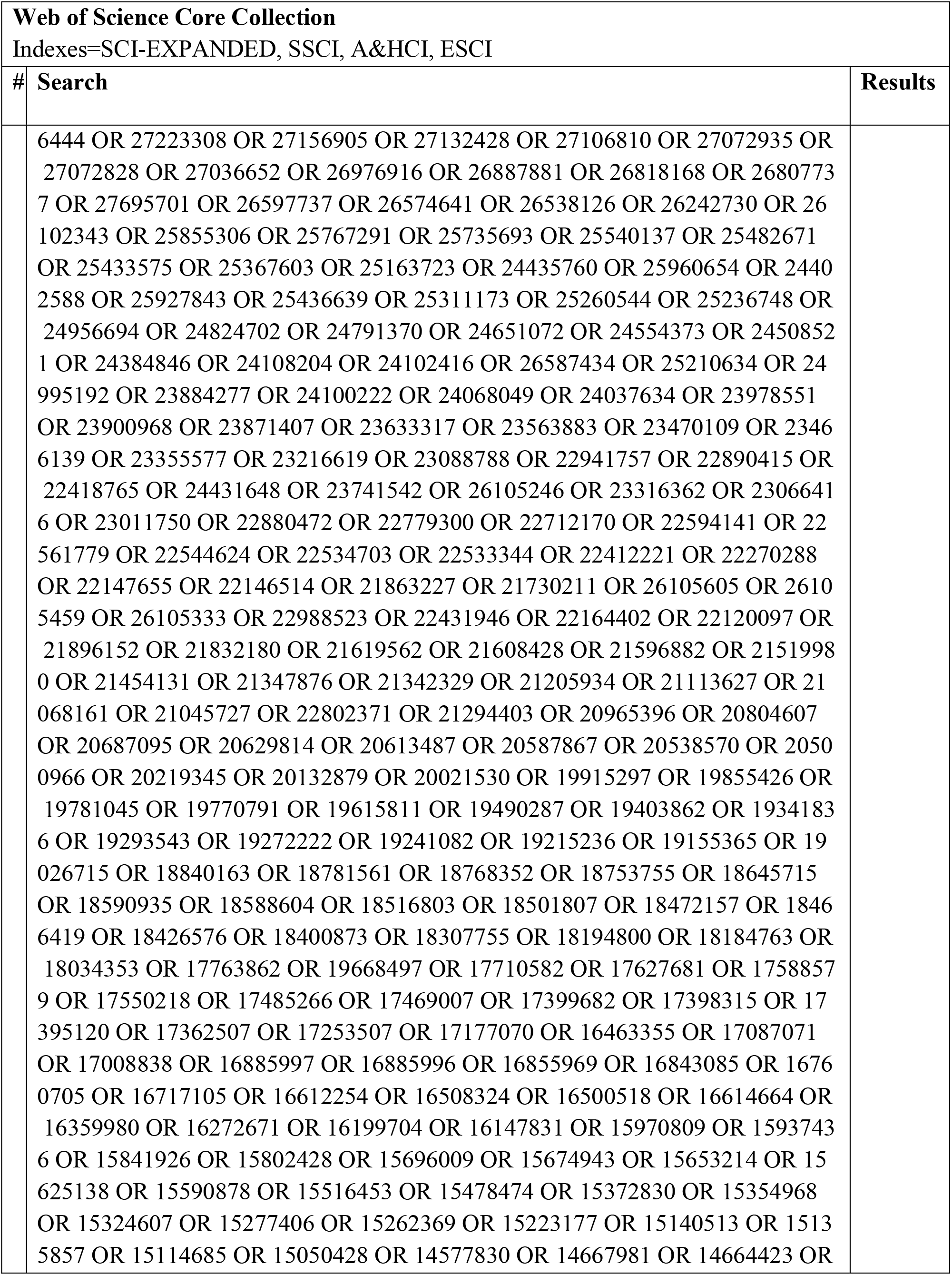
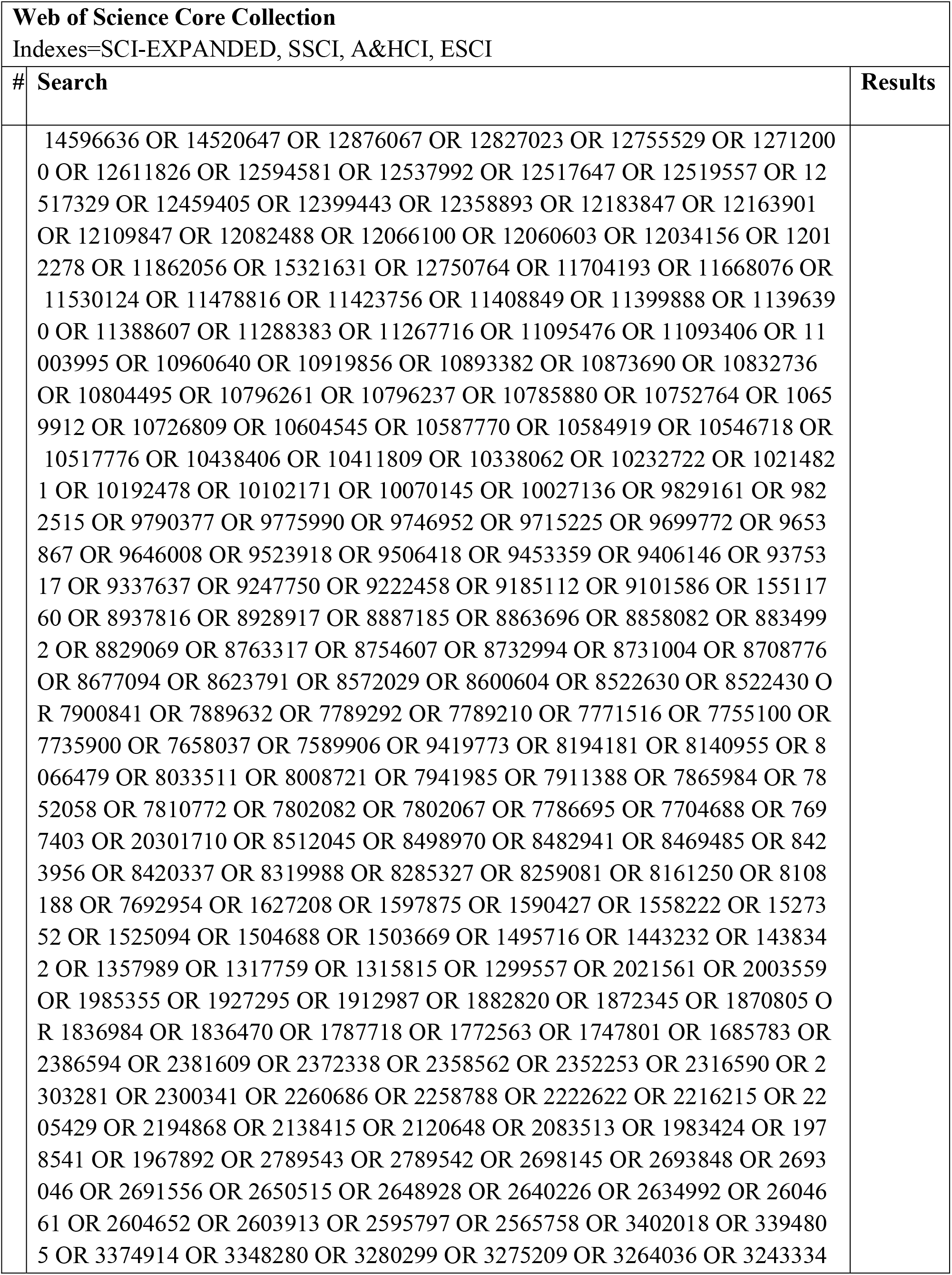
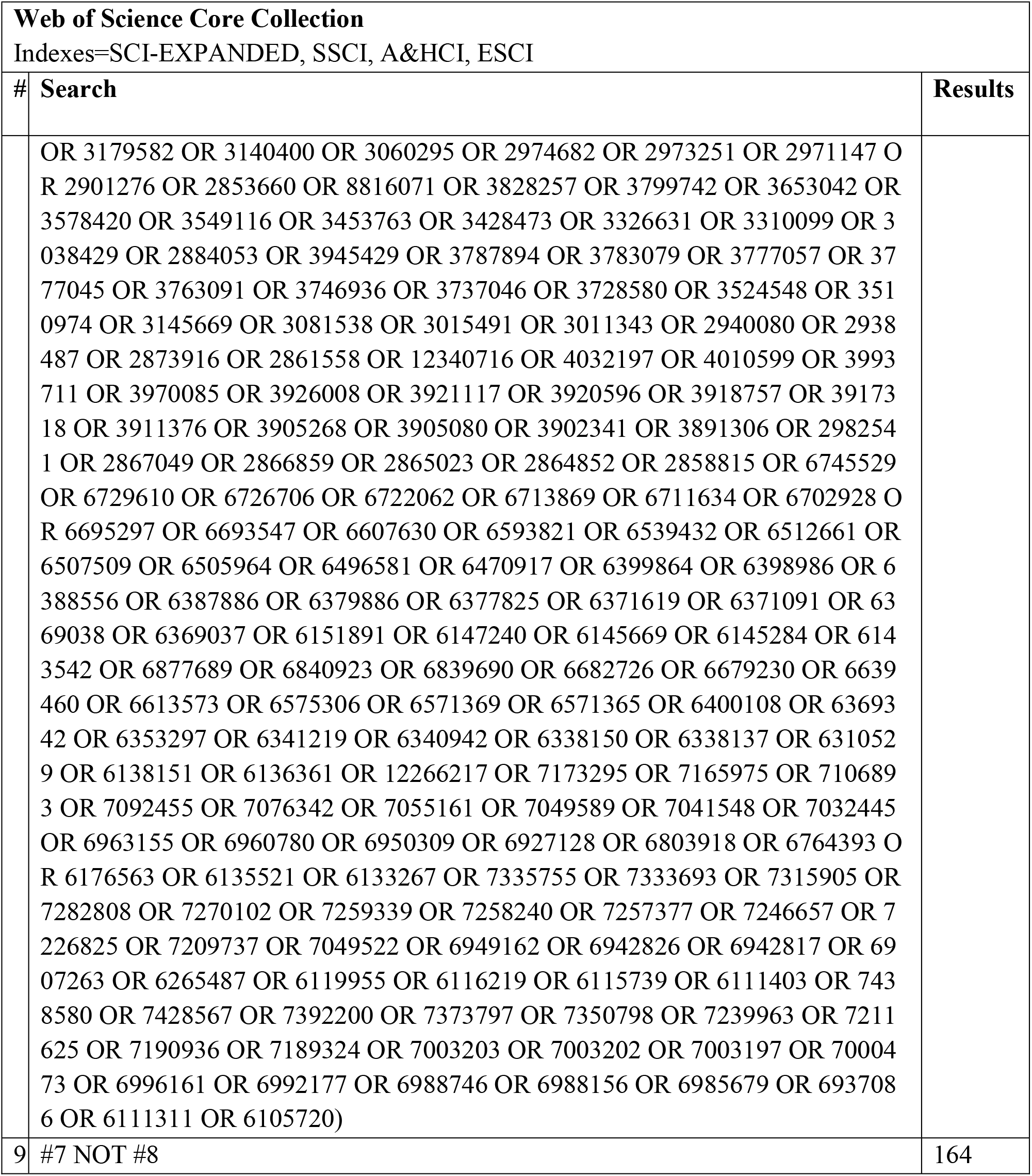

## Cochrane Central Register of Controlled Trials (CENTRAL)

| **Cochrane CENTRAL** |  |  |
| --- | --- | --- |
| # | Search | Results |
| #1 | MeSH descriptor: [HELLP Syndrome] this term only | 52 |
| #2 | MeSH descriptor: [Hypertension, Pregnancy-Induced] this term only | 205 |
| **#** | **Search** | **Results** |
| #3 | MeSH descriptor: [Pre-Eclampsia] this term only | 995 |
| #4 | MeSH descriptor: [Pregnancy Complications] this term only | 1736 |
| #5 | MeSH descriptor: [Puerperal Disorders] this term only | 309 |
| #6 | gestational hyperten*:ti,ab,kw | 1643 |
| #7 | hellp:ti,ab,kw | 217 |
| #8 | hypertensive disorders of pregnancy:ti,ab,kw | 257 |
| #9 | pre-eclamp*:ti,ab,kw | 1928 |
| #10 | preeclamp*:ti,ab,kw | 3537 |
| #11 | (postpartum NEAR/4 hyperten*):ti,ab,kw | 111 |
| #12 | (pregnan* NEAR/4 hyperten*):ti,ab,kw | 1849 |
| #13 | pregnancy-induced hyperten*:ti,ab,kw | 732 |
| #14 | toxaemia*:ti,ab,kw | 38 |
| #15 | toxemia*:ti,ab,kw | 376 |
| #16 | #1 OR #2 OR #3 OR #4 OR #5 OR #6 OR #7 OR #8 OR #9 OR #10 OR #11 OR #12 OR #13 OR #14 OR #15 | 7362 |
| #17 | MeSH descriptor: [Blood Pressure] this term only | 26829 |
| #18 | MeSH descriptor: [Postpartum Period] this term only | 1212 |
| #19 | #17 AND #18 | 24 |
| #20 | MeSH descriptor: [Pregnancy] this term only | 22025 |
| #21 | #17 AND #20 | 656 |
| #22 | MeSH descriptor: [Hypertension] this term only | 17835 |
| #23 | #22 AND #18 | 10 |
| #24 | #22 AND #20 | 353 |
| #25 | ((((arterial OR blood OR diastolic OR systolic) NEAR/1 pressur*) AND (post-partum OR postpartum OR puerper*)):ti,ab,kw | 11361 |
| #26 | ((bp OR dbp OR sdp) AND (perinatal OR postnatal OR post-partum OR postpartum OR puerper*)):ti,ab,kw | 238 |
| #27 | (hyperten* AND (perinatal OR post-partum OR postpartum OR puerper*)):ti,ab,kw | 1057 |
| #28 | #19 OR #21 OR #23 OR #24 OR #25 OR #26 OR #27 | 12620 |
| #29 | #16 OR #28 | 17859 |
| #30 | MeSH descriptor: [Bendroflumethiazide] this term only | 207 |
| #31 | MeSH descriptor: [Diazoxide] this term only | 79 |
| #32 | MeSH descriptor: [Diuresis] this term only | 565 |
| #33 | MeSH descriptor: [Diuretics] this term only | 2373 |
| #34 | MeSH descriptor: [Eplerenone] this term only | 218 |
| #35 | MeSH descriptor: [Furosemide] this term only | 1185 |
| **#** | **Search** | **Results** |
| #36 | MeSH descriptor: [Hydrochlorothiazide] this term only | 2055 |
| #37 | MeSH descriptor: [Metolazone] this term only | 39 |
| #38 | MeSH descriptor: [Mineralocorticoid Receptor Antagonists] this term only | 601 |
| #39 | MeSH descriptor: [Sodium Potassium Chloride Symporter Inhibitors] this term only | 113 |
| #40 | MeSH descriptor: [Spironolactone] this term only | 972 |
| #41 | MeSH descriptor: [Thiazides] this term only | 19 |
| #42 | MeSH descriptor: [Torsemide] this term only | 104 |
| #43 | Aldosterone Antagonist*:ti,ab,kw | 418 |
| #44 | Diazoxide:ti,ab,kw | 126 |
| #45 | diuresis:ti,ab,kw | 2156 |
| #46 | diuretic*:ti,ab,kw | 9331 |
| #47 | Eplerenone:ti,ab,kw | 490 |
| #48 | frusemide:ti,ab,kw | 355 |
| #49 | furosemide:ti,ab,kw | 2651 |
| #50 | hydrochlorothiazide*:ti,ab,kw | 3938 |
| #51 | lasix:ti,ab,kw | 77 |
| #52 | loop diuretic*:ti,ab,kw | 561 |
| #53 | metolazone:ti,ab,kw | 78 |
| #54 | Spironolactone:ti,ab,kw | 2079 |
| #55 | thiazide*:ti,ab,kw | 1391 |
| #56 | torsemide:ti,ab,kw | 141 |
| #57 | #30 OR #31 OR #32 OR #33 OR #34 OR #35 OR #36 OR #37 OR #38 OR #39 OR #40 OR #41 OR #42 OR #43 OR #44 OR #45 OR #46 OR #47 OR #48 OR #49 OR #50 OR #51 OR #52 OR #53 OR #54 OR #55 OR #56 | 16625 |
| #58 | #29 AND #57 | 135 |
| #59 | #29 AND #57 with Publication Year from 1980 to 2021, in Trials | 106 |

## Clinicaltrials.gov

Due to website constraints, each search was run separately using the “other terms” field. Results from each set were downloaded into Excel and deduplicated manually. The final 18 results were then added to EndNote to dedupe against all database results.

| **Clinicaltrials.gov** |  |  |
| --- | --- | --- |
| **#** | **Search** | **Results** |
| 1 | (Diuretics OR frusemide OR furosemide OR fursemide OR Hydrochlorothiazide OR lasix OR "loop diuretic" OR "Sodium Potassium Chloride Symporter Inhibitor" OR torsemide) AND Preeclampsia | 10 |
| 2 | (Diuretics OR frusemide OR furosemide OR fursemide OR Hydrochlorothiazide OR lasix OR "loop diuretic" OR "Sodium Potassium Chloride Symporter Inhibitor" OR torsemide) AND "hypertensive disorders of pregnancy" | 7 |
| 3 | (Diuretics OR frusemide OR furosemide OR fursemide OR Hydrochlorothiazide OR lasix OR "loop diuretic" OR "Sodium Potassium Chloride Symporter Inhibitor" OR torsemide) AND "hypertension, pregnancy-induced" | 12 |
| 4 | (Diuretics OR frusemide OR furosemide OR fursemide OR Hydrochlorothiazide OR lasix OR "loop diuretic" OR "Sodium Potassium Chloride Symporter Inhibitor" OR torsemide) AND "Gestational Hypertension" | 6 |

## REFERENCES

1. Hutcheon JA, Lisonkova S, Joseph KS. Epidemiology of pre-eclampsia and the other hypertensive disorders of pregnancy. Best Pract Res Clin Obstet Gynaecol. 2011;25:391–403

2. Ying W, Catov JM, Ouyang P. Hypertensive disorders of pregnancy and future maternal cardiovascular risk. J Am Heart Assoc. 2018;7:e009382

3. Clapp MA, Little SE, Zheng J, Robinson JN. A multi-state analysis of postpartum readmissions in the united states. Am J Obstet Gynecol. 2016;215:113.e111–113.e110

4. Hauspurg A, Jeyabalan A. Postpartum preeclampsia or eclampsia: Defining its place and management among the hypertensive disorders of pregnancy. Am J Obstet Gynecol. 2022;226:S1211–s1221

5. Kumar NR, Hirshberg A, Srinivas SK. Best practices for managing postpartum hypertension. Curr Obstet Gynecol Rep. 2022;11:159–168

6. Ouzounian JG, Elkayam U. Physiologic changes during normal pregnancy and delivery. Cardiol Clin. 2012;30:317–329

7. Malhamé I, Dong S, Syeda A, Ashraf R, Zipursky J, Horn D, et al. The use of loop diuretics in the context of hypertensive disorders of pregnancy: A systematic review and meta-analysis. J Hypertens. 2023;41:17–26

8. Lopes Perdigao J, Hirshberg A, Koelper N, Srinivas SK, Sammel MD, Levine LD. Postpartum blood pressure trends are impacted by race and bmi. Pregnancy Hypertens. 2020;20:14–18

9. Hauspurg A, Lemon L, Cabrera C, Javaid A, Binstock A, Quinn B, et al. Racial differences in postpartum blood pressure trajectories among women after a hypertensive disorder of pregnancy. JAMA Netw Open. 2020;3:e2030815

10. Sibai BM. Etiology and management of postpartum hypertension-preeclampsia. Am J Obstet Gynecol. 2012;206:470–475

11. Walters BN, Thompson ME, Lee A, de Swiet M. Blood pressure in the puerperium. Clin Sci (Lond). 1986;71:589–594

12. Magee L, von Dadelszen P. Prevention and treatment of postpartum hypertension. Cochrane Database Syst Rev. 2013:Cd004351

13. Bramer WM, Giustini D, de Jonge GB, Holland L, Bekhuis T. De-duplication of database search results for systematic reviews in endnote. J Med Libr Assoc. 2016;104:240–243

14. René Otten RdV, Linda Schoonmade. Amsterdam efficient deduplication (aed) method. 2019

15. Evidence Partners. Distillersr. 2021

16. Sterne JAC, Savović J, Page MJ, Elbers RG, Blencowe NS, Boutron I, et al. Rob 2: A revised tool for assessing risk of bias in randomised trials. Bmj. 2019;366:l4898

17. R Development Core Team. R: A language and environment for statistical computing. 2024

18. Ascarelli MH, Johnson V, McCreary H, Cushman J, May WL, Martin JN, Jr. Postpartum preeclampsia management with furosemide: A randomized clinical trial. Obstet Gynecol. 2005;105:29–33

19. Viteri OA, Alrais MA, Pedroza C, Hutchinson M, Chauhan SP, Blackwell SC, et al. Torsemide for prevention of persistent postpartum hypertension in women with preeclampsia: A randomized controlled trial. Obstet Gynecol. 2018;132:1185–1191

20. Lopes Perdigao J, Lewey J, Hirshberg A, Koelper N, Srinivas SK, Elovitz MA, et al. Furosemide for accelerated recovery of blood pressure postpartum in women with a hypertensive disorder of pregnancy: A randomized controlled trial. Hypertension. 2021;77:1517–1524

21. Siamansoori S, Afshari E, Palizdar M, Hosseini MA. Comparison of lasix and methyldopa in controlling hypertension in preeclampsia patients: A double-blind randomized clinical trial. Men’s Health Journal. 2020;4

22. Bozorgan TJ, Azadi P, Dehghani Z. Assessment of the effect of adding furosemide to antihypertensive treatment on postpartum hypertension in women with preeclampsia; a randomized clinical trial. Journal of Renal Injury. 2022

23. Dabaghi T, Shariati M, Laluha F, Movahhed F, Barikani A. Efficacy of postpartum furosemide therapy on blood pressure recovery in patients with severe preeclampsia: A randomized clinical trial. Bangladesh Journal of Medical Science. 2019;18:636–640

24. Veena P, Perivela L, Raghavan SS. Furosemide in postpartum management of severe preeclampsia: A randomized controlled trial. Hypertens Pregnancy. 2017;36:84–89

25. Matthews G, Gornall R, Saunders NJ. A randomised placebo controlled trial of loop diuretics in moderate/severe pre-eclampsia, following delivery. J Obstet Gynaecol. 1997;17:30–32

26. Cairns AE, Tucker KL, Leeson P, Mackillop LH, Santos M, Velardo C, et al. Self-management of postnatal hypertension: The snap-ht trial. Hypertension. 2018;72:425–432

27. Kitt JA, Fox RL, Cairns AE, Mollison J, Burchert HH, Kenworthy Y, et al. Short-term postpartum blood pressure self-management and long-term blood pressure control: A randomized controlled trial. Hypertension. 2021;78:469–479

28. Kitt J, Krasner S, Barr L, Frost A, Tucker K, Bateman PA, et al. Cardiac remodeling after hypertensive pregnancy following physician-optimized blood pressure self-management: The pop-ht randomized clinical trial imaging substudy. Circulation. 2024;149:529–541

29. Haas DM, Parker CB, Marsh DJ, Grobman WA, Ehrenthal DB, Greenland P, et al. Association of adverse pregnancy outcomes with hypertension 2 to 7 years postpartum. J Am Heart Assoc. 2019;8:e013092

30. Honigberg MC, Zekavat SM, Aragam K, Klarin D, Bhatt DL, Scott NS, et al. Long-term cardiovascular risk in women with hypertension during pregnancy. J Am Coll Cardiol. 2019;74:2743–2754

31. Stuart JJ, Tanz LJ, Rimm EB, Spiegelman D, Missmer SA, Mukamal KJ, et al. Cardiovascular risk factors mediate the long-term maternal risk associated with hypertensive disorders of pregnancy. J Am Coll Cardiol. 2022;79:1901–1913

32. Hauspurg A, Bryan S, Jeyabalan A, Davis EM, Hart R, Shirriel J, et al. Blood pressure trajectories through the first year postpartum in overweight or obese individuals following a hypertensive disorder of pregnancy. Hypertension. 2024;81:302–310

33. Emeruwa UN, Azad H, Ona S, Bejerano S, Alnafisee S, Emont J, et al. Lasix for the prevention of de novo postpartum hypertension: A randomized placebo-controlled trial (lapp trial). Am J Obstet Gynecol. 2024

34. Goel A, Maski MR, Bajracharya S, Wenger JB, Zhang D, Salahuddin S, et al. Epidemiology and mechanisms of de novo and persistent hypertension in the postpartum period. Circulation. 2015;132:1726–1733

